# The plasma proteins GSN, F2, CRTAC1, and HP reflect multigenerational health, longevity, and resilience

**DOI:** 10.64898/2026.02.06.26345744

**Authors:** Pasquale C. Putter, Marian Beekman, Nico Lakenberg, Jan-Wilm Lackmann, Stefan Müller, Roman-Ulrich Müller, Joris Deelen, P. Eline Slagboom, Philipp Antczak, Niels M.A. van den Berg

## Abstract

**Background:** The risk of chronic diseases and multimorbidity increases with age, yet, individuals of the same age can strongly differ in healthspan, ranging from early manifestation of age-related disease to robust health into very old age. Plasma biomarkers, including metabolites and proteins, can capture intrinsic health status, thereby providing insights into the nature of this variation. These biomarkers have been widely explored to understand chronic and early disease risk but less so for disease resilience in mid- and late-life survival (i.e. after 90 years), or for multigenerational longevity.

**Methods:** We quantified 326 plasma proteins using data-independent mass spectrometry in two generations of the Leiden Longevity Study cohort: F1 nonagenarian siblings (late-life; age ≥89; N=852) and F2 offspring and their partners (mid-life; age 30-80; N=2,282). Baseline plasma protein levels were tested for association with mid- and late-life survival, with up to 22 years of follow-up, and cardiometabolic healthspan, with up to 16 years of follow-up. By comparing F2 offspring and partners, we tested for plasma proteins associating with familial longevity.

**Findings:** Four proteins: GSN, F2, CRTAC1, and HP, consistently associated with increased mid- and late-life survival, prolonged cardiometabolic healthspan, and familial longevity; representing overall resilience. Moreover, six proteins: APCS, C7, FCN2, HPR, GSN, and PIGR, associated with mortality independent of MetaboHealth, a well-established metabolomics-based mortality score.

**Interpretation:** We identified GSN, F2, CRTAC1, and HP as promising candidate indicators of healthy aging and resilience, meriting further study.

**Funding:** ZonMw, LUF, BBMRI-NL, VOILA, Jörg Bernards-Stiftung, Köln Fortune, and CECAD

**Research in context:** *Evidence before this study:* As the population ages, implementing biomarkers that are able to distinguish vulnerable individuals from highly resilient ones may help to alleviate the burden on our healthcare system. Several studies have shown that plasma-derived proteins change with chronological age and can be used to discern those at risk of disease or early mortality. However, most plasma proteome studies to date have focused either on cross-sectional analyses, short follow-up periods, and/or cohorts with a narrow age range, often centered around mid-life. These studies therefore do not capture the biological factors that contribute to survival up to high ages (longevity), disease resilience and sustained health. In addition, it remains unclear whether the proteins and underlying mechanisms associated with increased survival are consistent across mid and late-life.

*Added value of this study:* This study set out to investigate the potential of plasma proteins as markers for healthy aging and resilience, and examines the extent to which these associations are consistent across generations. Using the family-based Leiden Longevity Study cohort, including long-lived nonagenarian siblings, their middle-aged offspring, and their partners, we leveraged a unique design to study survival, mid-life health, and multigenerational longevity. We show that prospective (longevity) survival is associated with both age-dependent and age-independent protein effects. Most age-independent proteins displaying consistent associations across life stages. In addition, relative protein level differences were detectable on average ten years before the onset of cardiometabolic disease in mid-life, indicating signatures of future resilience or vulnerability. Four proteins (GSN, CRTAC1, F2, and HP) show consistent association with survival-related outcomes and are proposed as candidate markers of healthy aging. We further observe favorable differences in these proteins in families enriched for longevity, which is indicative of lower disease risk. These identified proteins implicate inflammatory and osteoarthritis related processes. Furthermore, we demonstrate that proteins capture survival domains distinct from those reflected by metabolomic measures, demonstrating the complementary value of plasma proteomics.

*Implications of all the available evidence:* These findings highlight candidate protein biomarkers of healthy aging and resilience, which may provide improved insight into the mechanisms underlying healthy aging. They also facilitate the development of more sensitive indicators of health, whose implementation may enable earlier and more effective interventions in both population health and clinical care.

## Introduction

Currently around 65% of the middle-aged population (65+) in high-income countries is diagnosed with two or more chronic diseases (1). Cardiometabolic diseases represent a major part of this burden, with heart disease and stroke ranking among the leading sources of illness and mortality (2). The incidence of these diseases increases steadily from mid-life into older age (1). Yet, chronological age alone does not capture the wide variability of healthspan observed across individuals (3). Some individuals develop age-related diseases early, while others stay healthy into advanced age. This variation between individuals becomes more pronounced with age and as the population continues to age, the strain on healthcare systems is projected to increase significantly. Postponement of the onset of disease, discriminating vulnerable and resilient older adults, and increasing healthspan would contribute to reducing the strain on the healthcare system and ease societal challenges (1, 4). Discriminating the health status of older individuals and their physiological vulnerability, or resilience, is relevant for both population health and adequate clinical care. Molecular markers can assist in such discrimination and may provide insights into the biological mechanisms that underlie differences in health trajectories.

Over the past decade, plasma-derived molecular biomarkers have been developed using a range of omics technologies to quantify age-related physiological vulnerability and disease (5–7). Previously, we developed the MetaboHealth score, based on circulating metabolites, trained to predict future mortality (8). MetaboHealth has been shown to reflect health status in mid- and late-life, capturing underlying vulnerabilities such as frailty (5), cognitive decline (9), and hospitalization and survival in COVID-19 patients (10). Plasma proteomics provides a complementary dimension to metabolites by representing the immune system and tissue health through excreted proteins, which closely reflect disease states and constitute the majority of druggable targets in medicine (11). High-throughput proteomic platforms, such as OLINK or SomaScan, now allow for the simultaneous quantification of hundreds to thousands of plasma proteins through either direct mass spectrometry or affinity-based measurement methods.

To date, most proteomic studies on aging are performed in large cohort studies and are focused on identifying proteins, or protein signatures, associated with chronological age, disease risk, or vulnerability (7, 12–16), but not resilience across the lifespan. In addition, many of these studies are limited by cross-sectional designs, focus primarily on middle-aged individuals, and/or are constrained by short follow-up periods for health outcomes and mortality (17–22). As such, little is known about how comparable the association between plasma protein levels and low mortality risk in mid- and late-life are. In previous research, we showed that lifespan and healthspan are quantitatively linked to familial longevity; each additional long-lived family member contributing to greater lifespan and healthspan benefits in descendants (23, 24). Proteins associated with familial longevity may provide insights into resilience to physiological vulnerability and mortality in mid- and late-life. Longitudinal studies of such families may reveal which proteins reflect resilience across the life course, by association to cardiometabolic healthspan, low mid- and late-life mortality, and a strong familial longevity history. Identifying such proteins may reveal biomarkers capable of discriminating highly resilient from vulnerable individuals, with potential applications in both population health and geriatric care.

In this study, we employ data-independent (DIA) mass-spectrometry (MS)–based plasma proteomics to evaluate the potential of plasma proteins as robust indicators of healthy aging and resilience. We define these outcomes by focusing on: low mid- and late-life mortality, extended cardiometabolic healthspan, and a documented family history of longevity. These are investigated in data from the Leiden Longevity Study (LLS; **Supplementary Table 1**), including the late-life F1 nonagenarian siblings (age ≥ 89 years) and mid-life F2 generation (age 30-80 years; 25). Plasma proteins are tested for association with [1] all-cause mortality in mid- and late-life, with up to 22 years of mortality follow-up; [2] cardiometabolic healthspan in middle age, with up to 16 years of morbidity follow-up; [3] familial longevity, as quantified by the Longevity Relatives Count (LRC) score (26). We integrate these findings to identify robust candidate markers of healthy aging and resilience. Finally, we investigate how these proteins compare to our previously established vulnerability marker MetaboHealth and evaluate whether they capture similar or distinct mortality domains.

## Methods

### Study Design – the Leiden Longevity Study (LLS)

The LLS is a family-based study that comprises 421 families and was initiated in 2002 to study the mechanisms of healthy aging and exceptional survival. Inclusion took place between 2002 and 2006 and initially started with the recruitment of living sibling pairs (nonagenarian siblings, N=944). Within a sibling pair, males were invited to participate if they were 89 years or older and females if they were 91 years or older. Inclusion was subsequently extended to the children (F2 offspring, N=1671) of the nonagenarian siblings and their partners (F2 partners, N=744), which we collectively refer to as the F2 generation. In addition, demographic data of parents, non-included F1 siblings, and the F0 parents were linked to the nonagenarian siblings using genealogical records (**Supplementary Table 2**).

The LLS protocol was approved by the Medical Ethical Committee of the Leiden University Medical Center before the start of the study (P01.113) at 16^th^ of August 2002. The first participant was enrolled at 5^th^ of September 2002. In accordance with the Declaration of Helsinki, the LLS obtained informed consent from all participants prior to their entering the study.

### Mortality data

Up to 22 years of mortality information (until April 2024) is available within the LLS since the time of recruitment at 2002. Mortality information for the non-included family members was verified by birth or marriage certificates and passports whenever possible. Additional verification took place via personal cards which were obtained from the Dutch Central Bureau of Genealogy. In April 2024 mortality information for the nonagenarian siblings and F2 generation (including vital status and date of death) was updated through the Personal Records Database (PRD) which is managed by Dutch governmental service for identity information. https://www.government.nl/topics/personal-data/personal-records-database-brp. The combination of officially documented information provides very reliable and complete ancestral as well as current mortality information.

### Morbidity data

Healthspan data with up to 16 years of follow-up after study inclusion, was retrieved from the General Practitioners (GPs) of the LLS F2 generation and covers the period from birth until 2018. The morbidity history was available in a subset of the F2 generation participants (N=1,640; 71.9%). The GP records are kept up to date when a person switches from one GP practice to another. GPs were instructed to extract the presence of chronic age-related disease and the year the disease occurred from their electronic health records.

To increase statistical power for detecting proteins associated with cardiometabolic healthspan and to examine their associations with cardiometabolic disease (CMD) incidence, we clustered CMDs and defined them according to the International Statistical Classification of Diseases and Related Health Problems (ICD-10) codes. The following diseases were considered as a CMD: transient ischemic attack (TIA; ICD10: I60-I69), cerebrovascular accident (CVA; ICD10: I63), angina pectoris (AP; ICD10: I20), myocardial infarction (MI; ICD10: I21), Hypertension (ICD10: I10), Diabetes (ICD10: E10-14).

### Longevity Relatives Count (LRC)-score

The Longevity Relatives Count (LRC) score was used to assess the family history of longevity in the F2 generation (**Formula 1**; (26). The LRC score indicates the proportion of ancestors that became long-lived, defined as belonging to the top 10% survivors of their birth cohort, as calculated by the lifetables of Statistic Netherlands (27). The LRC is weighted by the genetic distance between partners/offspring and their ancestors. For example, an LRC of 0.5 indicates 50% long-lived ancestors. For this study, two generations of ancestors were available to calculate the LRC score for the offspring and one generation for the partners (**Supplementary Table 3** for the LRC distribution).

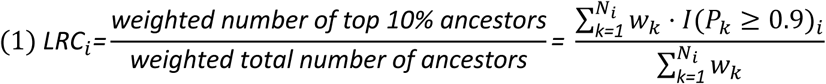

Where *i* refers to the participant for whom the score is built. *k* is an index referring to each ancestral blood relative of person *i* who are used to construct the score. *N*_*i*_ refers to the total number of ancestors of person *i*, *P*_*k*_ is the sex and birth year-specific survival percentile, based on lifetables, of ancestor *k*, and *I*(*P*_*k*_ ≥ 0.9)_*i*_indicates if ancestor *k* belongs to the top 10% survivors. 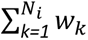 is the weighted total number of ancestors of participant *i*. The relationship coefficients are used as weights *w*_*k*_.

### ^1^H-NMR metabolomics

From EDTA plasma samples at time of recruitment, metabolomics data was quantified using high-throughput nuclear magnetic resonance (^1^H-NMR) spectroscopy by the Nightingale Health platform in 2014. MetaboHealth, is a 14 metabolite-based marker trained on 10-year all-cause mortality that was also shown to reflect current health and frailty status (5, 8). The Nightingale Health platform contains over 250 metabolomic measurements, of which we focus on the 14 metabolites comprising MetaboHealth (Acetoacetate (AcAce), Albumin (Alb), Glucose (Glc), Glycoprotein acetyls (Gp), Histidine (His), Isoleucine (Ile), Lactate (Lac), Leucine (Leu), Phenylalanine (Phe), the Ratio of polyunsaturated fatty acids to total fatty acids (PUFA/FA), Total lipids in small HDL (S-HDL-L), Valine (Val), Mean diameter for VLDL particles (VLDL-D), and Total lipids in chylomicrons and extremely large VLDL (XXL-VLDL-L)). Quality control followed the methodology outlined in the original Metabohealth paper (8). After excluding outlier samples and those with high missingness, in both datasets separate each metabolite was Rank-Inversed Normalized (RIN) and Z-scaled to have a standard deviation (SD)±1 and mean = 0. The MetaboHealth weighted sumscore was then calculated using the Beta-coefficients from the original paper (8) (**Formula 2; Supplementary Table 4**).

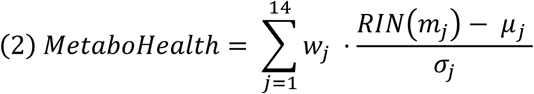

Where *m*_*j*_are the raw quantified values of metabolite *j* after quality control. *RIN*(*m*_*j*_) denotes the rank inverse normalized values of *m*_*j*_. *μ*_*j*_ and *σ*_*j*_ are the mean and standard deviation of *RIN*(*m*_*j*_) in each dataset. *w*_*j*_is the weight assigned to metabolite *j*, which correspond to the beta’s derived from the original MetaboHealth publication (8); **Supplementary Table 3**)

### Plasma proteomics using data-independent mass spectrometry acquisition

#### Plasma proteome measurements

Sodium citrate plasma samples from time of recruitment, were prepared in 2023 following a modified SP3 protocol performed on an Integra Assist plus (Integra), foregoing the peptide cleanup on the second day (28). Beads were removed after acidification and samples cleaned by using mixed-mode StageTips (29). Samples were analyzed on an UltiMate 3000 coupled to an Orbitrap Exploris 480 with FAIMS pro (all Thermo Scientific).

Next, samples were loaded onto a precolumn (PepMap 5 mm cartridge, Thermo Scientific) and reverse flushed onto an in-house packed 30 cm pulled tip column (150 µm inner diameter, filled with 2.7 µm Poroshell EC120 C18, Agilent). Separation took place on a 25 min gradient running 0.1 % formic acid (eluent A) against 80 % acetonitrile, 0.1 % formic acid (eluent B). Gradient started at 6 % B and increased to 35 % over 22 min followed by washing and equilibration to standard conditions, all with a constant flow of 1 µl/min. FAIMS set to -50 V compensation voltage with inner and outer electrode temperature kept constant at 99.5 °C and 85 °C, respectively. The mass spectrometer resolution of both MS1 and MS2 were set to 15k resolution and was running in data independent acquisition mode an 30 % normalized collision energy. The mass range from 380 to 900 m/z was covered in 30 staggered windows of 16 m/z each, resulting in effectively 8 m/z windows after deconvolution using ProteoWizard (30).

For library generation, a pool was generated from all samples and high pH fractionated on an 1 h gradient using an Infinity 1260 LC (Agilent). Resulting 96 fractions were concatenated into a total of 24 samples and analyzed on the identical setup and LC gradient used for sample analysis but with the mass spectrometer operated in Top 18 DDA. MS1 resolution was set to 60k, MS2 resolution to 30k with a dynamic exclusion of 20 s and an isolation window of 1.2 Th. The library was afterwards build using Fragpipe 15.0 and its predefined library building workflow with standard parameters and the Human Uniprot reference proteome including isoforms (downloaded 13.01.2021). The resulting library contained 8781 precursors from 681 proteins. Finally, samples were searched against the library using DIA-NN v1.8.1 with reannotation activated and the additional command line “—report-lib-info” (31).

#### Plasma Proteome Data Preprocessing

Raw MS spectra were processed using DIA-NN v1.8.1 and filtered to ensure that each identified protein had at least 1 unique peptide identifying it. In total N=3,373 samples were measured across 40 plates in 3 separate batches (**Supplementary Table 5A**). In total N=460 proteins were quantified (**Supplementary Table 5B**), proteins that had more than 50% missing data were removed, resulting a total of N=326 robustly quantified proteins, with Albumin being the most abundant and CD109 the least abundant (**Supplementary Table 5C**), the proteins in this protein panel were largely uncorrelated together (**Supplementary Table 5D**). No proteins were identified to be absent in either of the two groups in statistically significant numbers due to age effects. Outlier and low-quality samples were removed (N=61) from the analysis, as well as the pooled calibration samples (N=314), and distant LLS family members, such as uncles, aunts, and nephews, (N=264), resulting in a total of N=3,134 samples going into the analyses.

Missing values were imputed using a random normal distribution with a mean equal to the mean of the 5% proteins with the lowest intensities minus 1.6*log_2_ units. This ensured that all missing values were imputed just below the observed intensity levels. After, in each dataset (nonagenarian siblings and F2 generation) the proteins were Rank-Inversed Normalized (RIN) transformed and scaled to have a standard deviation (SD)±1 and mean = 0.

### Statistical analyses

Statistical analyses were conducted using R (v4.2.2). We reported 95% confidence intervals (CI) and considered a p-value of 0.05 as nominal statistically significant. Additionally, we controlled for multiple testing by applying a Benjamini-Hochberg False Discovery Rate (FDR) threshold of 0.1. Lme4 and Lmertest were used to fit the linear mixed models (LMM), and FrailtyEM for the Cox-type frailty (random effect) survival models (32). A random effect (LLM=gaussian distribution and Cox=Gamma distribution) was used to adjust for similarity among family members. For the proteomic analyses the Batch-ID was incorporated as a covariate in the model to account for technical variation between batches.

#### Survival analyses (Cox-type random effect models)

To investigate which proteins associate with survival in late-life (nonagenarian siblings), and mid-life (F2 generation), respectively, we fitted a set of univariate Cox-type random effect models:

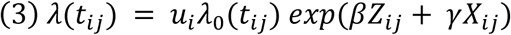

Here *t*_*ij*_ is the right-censored outcome of participant *j* in family *i*, *λ*_0_(*t*_*ij*_) refers to the general risk of death at a given age, and *β* represents the main regression coefficient of interest, representing the fixed effect of variable *Z* which indicates the protein of interest. The models are adjusted for left truncation given by the age at recruitment in the study and adjusted for the confounders Sex, and batch effect of the MS protein run denoted by *X*. *u*_*i*_ refers to an unobserved heterogeneity and random effect (frailty) shared by the members of the same family *i*, and was assumed to follow a gamma distribution.

To test the performance of the metabolomic-based vulnerability marker MetaboHealth in our cohort, with the newest mortality data, in the nonagenarian siblings and F2 generation. We fitted two Cox-type random effect models. Here *t*_*ij*_is the right-censored outcome of participant *j* in family *i*, *λ*_0_(*t*_*ij*_) refers to the general risk of death at a given age, and *β* represents the main regression coefficient of interest, representing the fixed effect of variable *Z* which indicates the MetaboHealth, which follows a normal distribution. The models are adjusted for left truncation given by the age at recruitment in the study and adjusted for the confounders Sex denoted by *X*. *u*_*i*_refers to an unobserved heterogeneity and random effect (frailty) shared by the members of the same family *i*, and was assumed to follow a gamma distribution.

To investigate which proteins associate with mortality independent of MetaboHealth in the nonagenarian siblings and F2 generation, we fitted a set of univariate Cox-type random effect models. To the base-model of the mortality analyses we added MetaboHealth, which follows a normal distribution, as a confounder denoted by *X*. *β* represents the main regression coefficient of interest, representing the fixed effect of variable Z which indicates the protein of interest adjusted for MetaboHealth.

To see which protein levels associate with cardiometabolic healthspan we assessed time till first CMD in our F2 generation, we focused on the subset in which we had disease data available (N=1,640; 71.9%) and were disease-free at baseline (N=1,235; 75.3%). In this group we fitted a set of univariate Cox-type random effect models. Here *t*_*ij*_is the right-censored outcome of participant *j* in family *i*, *λ*_0_(*t*_*ij*_) refers to the general risk of developing a CMD at a given age, and *β* represents the main regression coefficient of interest, representing the fixed effect of variable *Z* which indicates the protein of interest. The models are adjusted for left truncation given by the age at recruitment in the study (minus one year to ensure CMD incidence didn’t fall before the recruitment) and adjusted for the confounders Sex, and technical batch effect of the MS protein run denoted by *X*. *u*_*i*_ refers to an unobserved heterogeneity and random effect (frailty) shared by the members of the same family *i*, and was assumed to follow a gamma distribution.

#### Linear mixed-models

To investigate which proteins associate with familial longevity as quantified by the LRC-score in our F2 generation we fitted a set of univariate linear mixed-models:

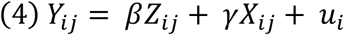

Where *Y*_*ij*_ is the protein level for person *j* in family *i*. *β* represents the main regression coefficient of interest, representing the fixed effect of variable *Z* which is the LRC-score for our F2 generation, the LRC-score follows a bi-model distribution (**Supplementary Table 3**). *γ* is a vector of regression coefficients for the fixed effects of possible confounders which are: age at recruitment, Sex, and technical batch effect of the MS protein run denoted by *X*. *u*_*i*_ refers to an unobserved heterogeneity and random effect (frailty) shared by the members of the same family *i*, and was assumed to follow a normal distribution.

## Results

### Population characteristics of the Leiden Longevity Study (LLS)

We quantified 326 proteins from citrate plasma samples collected at the time of recruitment and used mass-spectrometry in two generations from the family-based LLS (**Supplementary Table 1**), namely the nonagenarian siblings (N=852) as well as the F2 generation (N=2,282) (**Supplementary Table 5, 6**). In addition, we performed ^1^H-NMR metabolomics profiling in baseline EDTA plasma samples from the nonagenarian siblings (N=838; 98.4%) and F2generation (N=2,222; 97.4%), in which we calculated MetaboHealth (**Table 1**). Mortality follow-up extended up to 22 years from time of recruitment. By the beginning of 2024, all nonagenarian siblings had died (mean age at death = 97.4 years; SD ± 3.6) and in the F2 generation 20.3% (N=461) of the individuals had passed away (mean age at death = 76.4 years; SD ± 8.6). Disease data, with up to 16 years of follow-up, was available in a subset of the F2 generation (N=1,640; 71.9%). At time of recruitment, 75.3% (N=1,235) of the individuals were disease free, of those 39.8% (N=491) developed their first CMD after 10.1 years (SD ± 3.6). An overview of the study can be found in **Supplementary Table 1**.

**Table 1.** Characteristics of mortality and health in the two generations of the Leiden Longevity Study (LLS)

|  |  | Nonagenarian siblings | F2 generation |
| --- | --- | --- | --- |
| Total participants* | N | 852 | 2,282 |
| Sibships | N | 409 | 1,323 |
| Female | N (%) | 522 (61.3%) | 1,244 (54.5%) |
| Age at recruitment | mean (range) SD | 93.3 (88.9-103.4) ± 2.6 | 59.2 (30.2-80.3) ± 6.8 |
| Deaths<sup>†</sup> | N (%) | 851 (100%)<sup>1</sup> | 461 (20.3%)<sup>2</sup> |
| Age at death<sup>†</sup> | mean (range) SD | 97.4 (89.1-109.9) ± 3.6 | 76.4 (48.3-96.4) ± 8.6 |
| LRC-score | mean (range) SD | - | 0.34 (0-1) ± 0.2<sup>3</sup> |
| Disease data available<sup>‡</sup> | N (%) | - | 1,640 (71.9%) |
| Disease free at recruitment | N (%) | - | 1,235 (75.3%)<sup>4</sup> |
| Cardiometabolic disease incidence events<sup>‡</sup> | N (%) | - | 491 (39.8%)<sup>5</sup> |
| Plasma proteome | N (%) | 852 (100%) | 2,282 (100%) |
| MetaboHealth | N (%) | 838 (98.4%) | 2,222 (97.4%) |

N (%) is percentage of total participant group, unless stated as otherwise.
\*Total participants with blood proteome data available that met QC
† Mortality data available until April 2024
‡ General Practitioner health record disease data available from birth until 2018
<sup>1</sup> Lost to follow-up: 1
<sup>2</sup> Lost to follow-up: 9
<sup>3</sup> Distribution plot of the Longevity Relatives Count (LRC)-score shown in **Supplementary Table 3**
<sup>4</sup> % indicates disease free at recruitment from those in which we have disease data available
<sup>5</sup> % indicates number of cardiometabolic disease incidence events in those that were disease free at recruitment

### Proteins associated with survival in mid- and late-life

To establish which protein represents increased survival in late-life, we examined the association between the proteins and all-cause mortality in the nonagenarian siblings by a frailty (mixed-model) survival analysis. We identified 102 proteins which were nominally significantly (P≤0.05) associated with time to death (mortality; **Supplementary Table 7, 9**). Out of these 102 proteins, 89 remained significant after multiple testing correction (FDR≤0.1; **Figure 2A**). For 52 out of the 89 proteins, higher levels were associated with increased survival. The strongest effect was observed for GSN with a Hazard Ratio of 0.77 (HR_F1_ 0.77 [95% CI:0.71-0.84]). This indicates a strong survival advantage, as for every standard deviation increase in GSN protein levels the yearly risk of dying decreases with 23%. In addition, higher levels of SERPINA5 (HR_F1_ 0.80 [95% CI:0.74-0.87]) and SERPINA4 (HR_F1_ 0,81 [95% CI: 0.75-0.87]) were associated with a strong survival advantage. For the other 37 proteins higher levels were associated with a decreased survival advantage. The strongest effects were observed for C7 (HR_F1_ 1.31 [95% CI: 1.21-1.42]), C9 (HR_F1_ 1.26 [95% CI:1.17-1.36]), and SERPINA1 (HR_F1_ 1.22 [95% CI: 1.13-1.32]).

**Figure 2.**
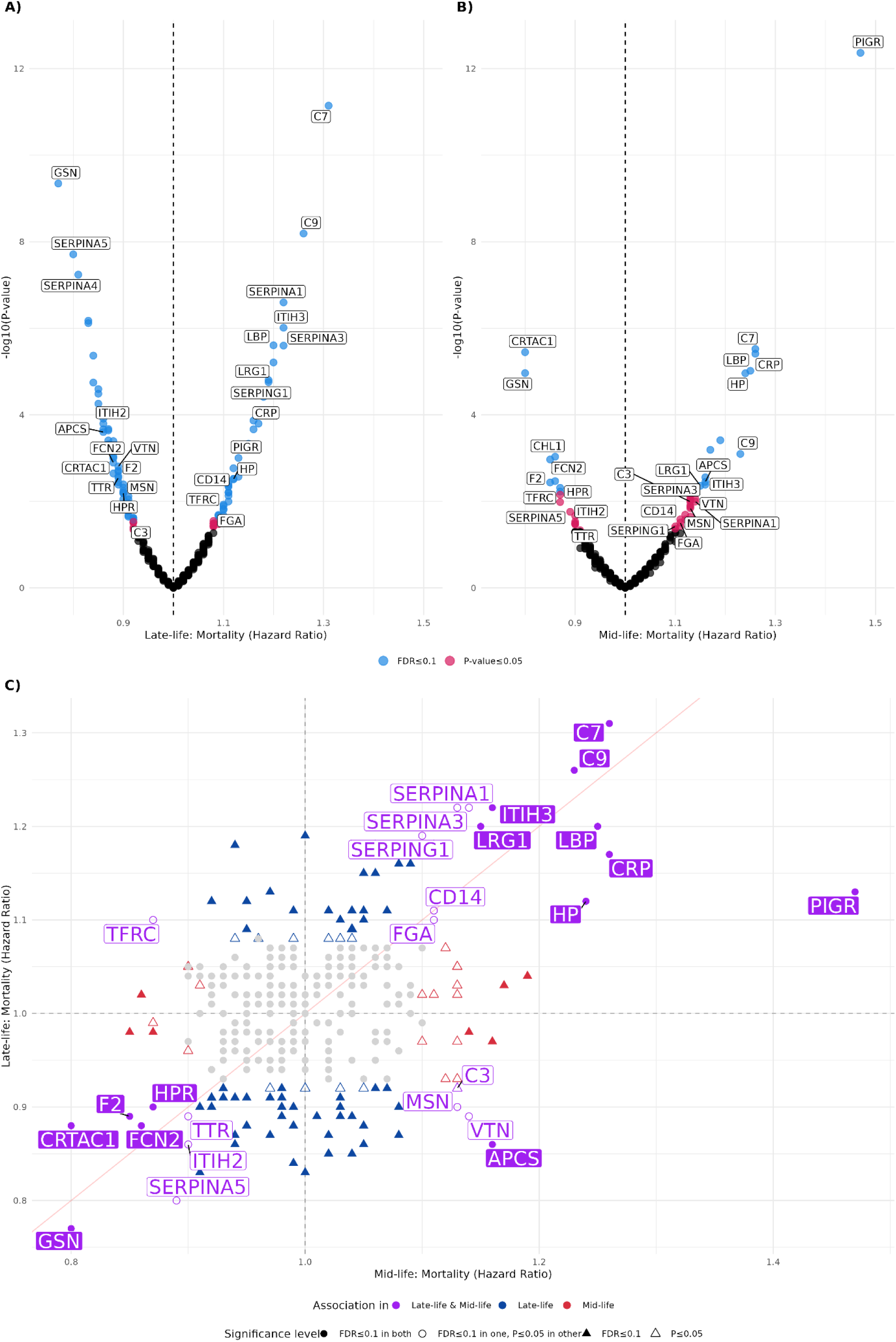
Prospective Plasma Protein Associations with survival in Mid- and Late-life. Prospective associations between 326 plasma proteins and survival as defined by all-cause mortality. (**A**) Late-life analysis in the nonagenarian siblings (males ≥89, females ≥91; mean age 93.3; N=852) with up to 18 years of follow-up. Proteins nominally significant proteins (P ≤ 0.05) are shown in red (n = 13), significant after FDR correction (≤0.1) are shown in blue (n = 89), and. (**B**) Mid-life analysis in the F2 generation (age 30–80; mean age 59.2; N=2,282) with up to 22 years of follow-up. Proteins nominally significant are shown in red (n = 28), significant after FDR correction (≤0.1) are blue (n = 21). Labeled proteins in panels A and B represent the strongest associations and those overlapping associations across cohorts. (**C**) Hazard–hazard plot comparing mid- and late-life associations; proteins nominally associated with mortality are labelled in purple (n = 26), and those significant after FDR correction (≤0.1) are filled in purple (n = 14). Detailed results are provided in **Supplementary Table 7,9.**

To investigate whether associations of plasma proteins with mortality have an age-dependent effect we with repeated our analyses for all-cause mortality in mid-life. To this end, we performed a frailty (mixed-model) survival analysis in the F2 generation. We identified 49 proteins which were nominally significantly associated with mid-life mortality (**Supplementary Table 7, 9**). Out of the 49 proteins, 21 remaining significant after multiple testing correction (**Figure 2B**). Among these, higher levels of 8 proteins were associated with an increased survival, while higher levels of 13 proteins were associated with decreased survival. The proteins with the strongest survival advantage included CRTAC1 (HR_F2_ 0.80 [95% CI: 0.73-0.89]), GSN (HR_F2_: 0.80 [95% CI: 0.72-0.88]), and CHL1 (HR_F2_ 0.88 [95% CI:0.78-0.94]), while those with the strongest survival decrease included PIGR (HR_F2_ 1.47 [95% CI: 1.33-1.62]), C7 (HR_F2_ 1.26 [95% CI: 1.14-1.39]), and CRP (HR_F2_ 1.26 [95% CI: 1.14-1.39]).

When comparing the mortality associations between mid- and late-life for age-independent associations we identified 26 mortality-associated proteins that were nominally significant in both the F2 generation and the nonagenarian siblings (**Figure 2C; Purple; Supplementary Table 9**). Among these age-independent associations, five proteins exhibited opposite effects in mid- and late-life. Four proteins (APCS, VTN, MSN, and C3) showed an decreased survival in mid-life but an increased survival in late-life with increasing protein levels. In contrast, one protein (TFRC) displayed the opposite trend, being associated with increased survival in mid-life and decreased survival in late-life. The other 21 protein associations are concordant in their relation between mid- and late-life risk. Of these 21, 13 proteins remained significant after multiple testing correction in the nonagenarian siblings and F2 generation. For five proteins (GSN, CRTAC1, F2, FCN2, and HPR) higher levels were associated with an increased survival as their levels increased, while for the other eight (C7, C9, ITIH3, LRG1, LBP, CRP, HP, and PIGR) higher levels were associated with decreased survival.

A subset of proteins demonstrated age-dependent associations with survival. In the nonagenarian siblings, 75 proteins showed an association solely with survival in late-life (**Figure 2C; Blue; Supplementary Table 9**), among these 64 remained significant after multiple testing. For 23 of these proteins higher levels were associated with increased survival at late-life, while for the other 41 proteins higher levels were associated with decreased survival in late-life. The strongest survival advantage was observed for FCN3, ITIH1, and SERPINA4, while B2M, CFD, and CP showed the strongest late-life mortality risk. In the F2 generation 23 proteins showed an association only with mid-life survival (**Figure 2C; Red; Supplementary Table 9**), of which seven remained significant after multiple testing. For three proteins (CHL1, HSPG2, and IGKV2D-30) higher levels were associated with increased survival in mid-life, while for the other four proteins (CFB, KLKB1, LGALS3BP, and SAA4) higher levels were associated with decreased survival in mid-life.

Together these findings suggest that mortality-risk is associated with both age-dependent and age-independent proteins. Among the proteins with age-independent associations, most show consistent effects across mid- and late-life, though the strength of each of these effects varies. However, a small subset of proteins exhibits crossover effects, i.e. shows opposite associations with mortality risk between life stages.

### Protein levels associate with prolonged cardiometabolic healthspan

To assess the potential of the proteins as biomarkers capable of discriminating highly resilient from vulnerable individuals in mid-life, we examined disease-free individuals at baseline and assessed their cardiometabolic healthspan, defined as the time to first cardiometabolic disease (CMD), as diagnosed from GP records. This analysis was limited to middle age, as healthspan data was not available for the late-life nonagenarian siblings. We conducted a frailty (mixed-model) survival analysis using the disease-free baseline subset of the F2 generation (N=1,235; **Table 1**). We observed that 45 proteins were nominally significantly associated with the risk of developing a first CMD (**Supplementary Table 7, 9**), of which 15 proteins remained significant after FDR correction (**Figure 3**). Out of the 15 proteins, higher levels of 5 proteins were associated with a decreased CMD risk, while higher levels of 10 proteins were associated with an increased CMD risk. GSN (HR 0.74 [95% CI: 0.66-0.84]), DPP4 (HR 0.80 [95% CI: 0.72-0.90]), and AMBP (HR 0.82 [95% CI: 0.73-0.93]) showed the strongest positive associations with cardiometabolic healthspan, while LGALS3BP (HR 1.29 [95% CI: 1.14-1.45]), SAA4 (HR 1.26 [95% CI: 1.12-1.41]), and CRP (HR 1.21 [95% CI: 1.08-1.36]) showed the strongest negative associations. These results highlight a select subset of proteins as potential biomarkers for CMD incidence and prolonged cardiometabolic healthspan, with relative detectable level differences in the plasma with on average ten years before disease onset.

**Figure 3.**
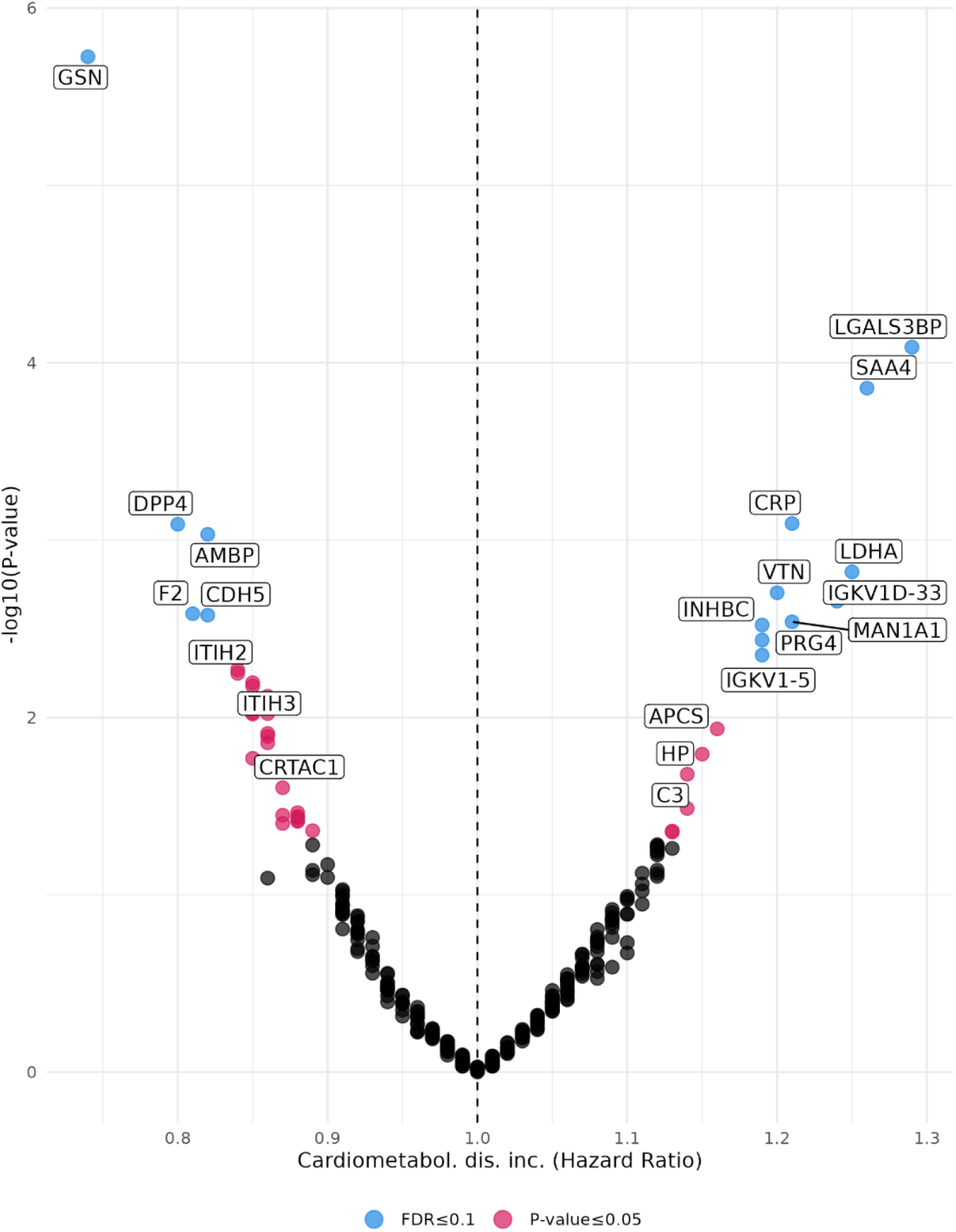
Prospective Plasma Protein Association with Cardiometabolic Healthspan at Mid-life. Volcano plot showing associations between 326 plasma proteins and cardiometabolic healthspan as defined as incident of cardiometabolic disease (transient ischemic attack, cerebrovascular accident, angina pectoris, myocardial infarction, hypertension, or diabetes) at mid-life (age 30–80 at recruitment; mean age 59.2) in the F2 generation who were disease-free at baseline (n=1,235), with up to 16 years of follow-up. Proteins significantly associated with disease after FDR correction (≤0.1) are shown in blue (n = 15), and nominally significant proteins (P ≤ 0.05) are shown in red (n = 30). Detailed results are provided in **Supplementary Table 7,9**. Labeled proteins represent the FDR associated proteins and those that were identified to be associated with mid- and late-life survival.

### Plasma proteome associations with familial longevity

In our previous work, we developed the Longevity Relatives Count (LRC) score to quantify familial clustering of longevity (26). Individuals with high LRC-scores not only had increased survival advantages across the entire life course but also showed increased cardiometabolic healthspan, for over a decade later than their low-scoring partners (24). Thus, we investigated if these survival advantages are reflected in the plasma proteome of individuals with ancestral longevity, as quantified with the LRC score, using mixed-model regression analysis in the F2 generation. 31 proteins were nominally significantly associated with the LRC-score (**Supplementary Table 8, Table 2**). Three proteins remained significant after multiple testing correction: CPN1 (β 0.4 [95% CI:0.2-0.6]), GPX3 (β 0.38 [95% CI:0.18-0.58]), and SHBG (β 0.35 [95% CI: 0.15-0.55]). For all three proteins, higher protein levels associated with higher levels of familial longevity.

**Table 2.**
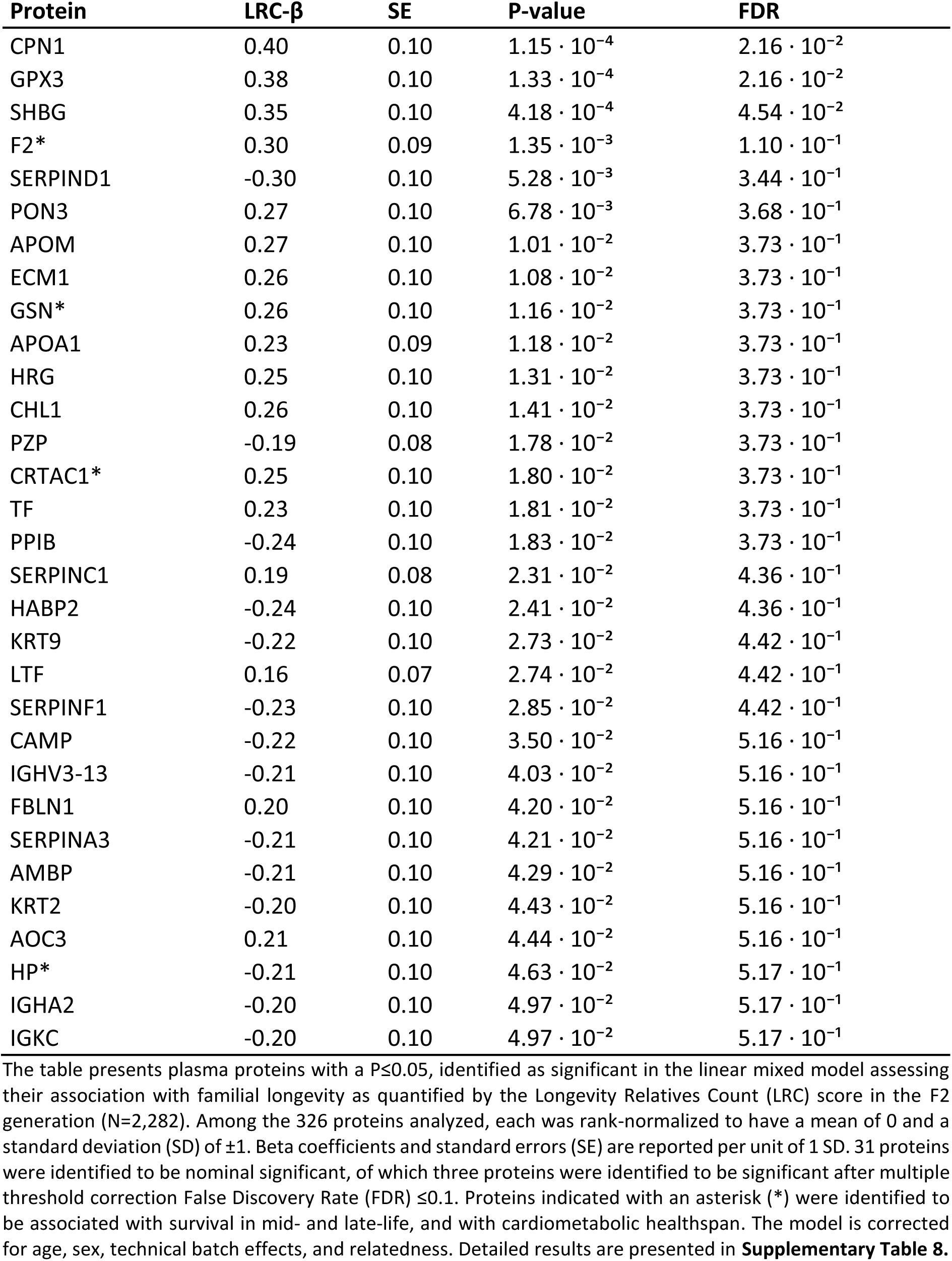
Plasma Proteins Associated with Familial Longevity.

### Candidate Protein Markers of Healthy Aging and Resilience

To identify proteins that may be potential indicators of healthy aging, we identified overall resilience proteins which associated with survival in mid- and late-life, cardiometabolic healthspan, and familial longevity (**Figure 4**). Ten proteins (GSN, CRTAC1, F2, HP, CRP, ITIH2, ITIH3, APCS, VTN, and C3) were nominally significant across mid- to late-life and cardiometabolic healthspan. Moreover, four of these proteins (GSN, CRTAC1, F2, and HP) were also significantly associated with familial longevity, making them candidate resilience makers of healthy aging in mid- and late-life. Higher levels of GSN, CRTAC1, and F2 were associated with increased survival across both life stages, and to a prolonged cardiometabolic healthspan. Additionally, these protein levels were higher with increasing familial longevity scores, consistent with a resilience profile. In contrast, higher HP levels were indicative of frailty as it associated with decreased survival and cardiometabolic healthspan. Whereas increasing levels of familial longevity scores corresponded to lower HP levels, consistent with an increased resilience profile.

**Figure 4.**
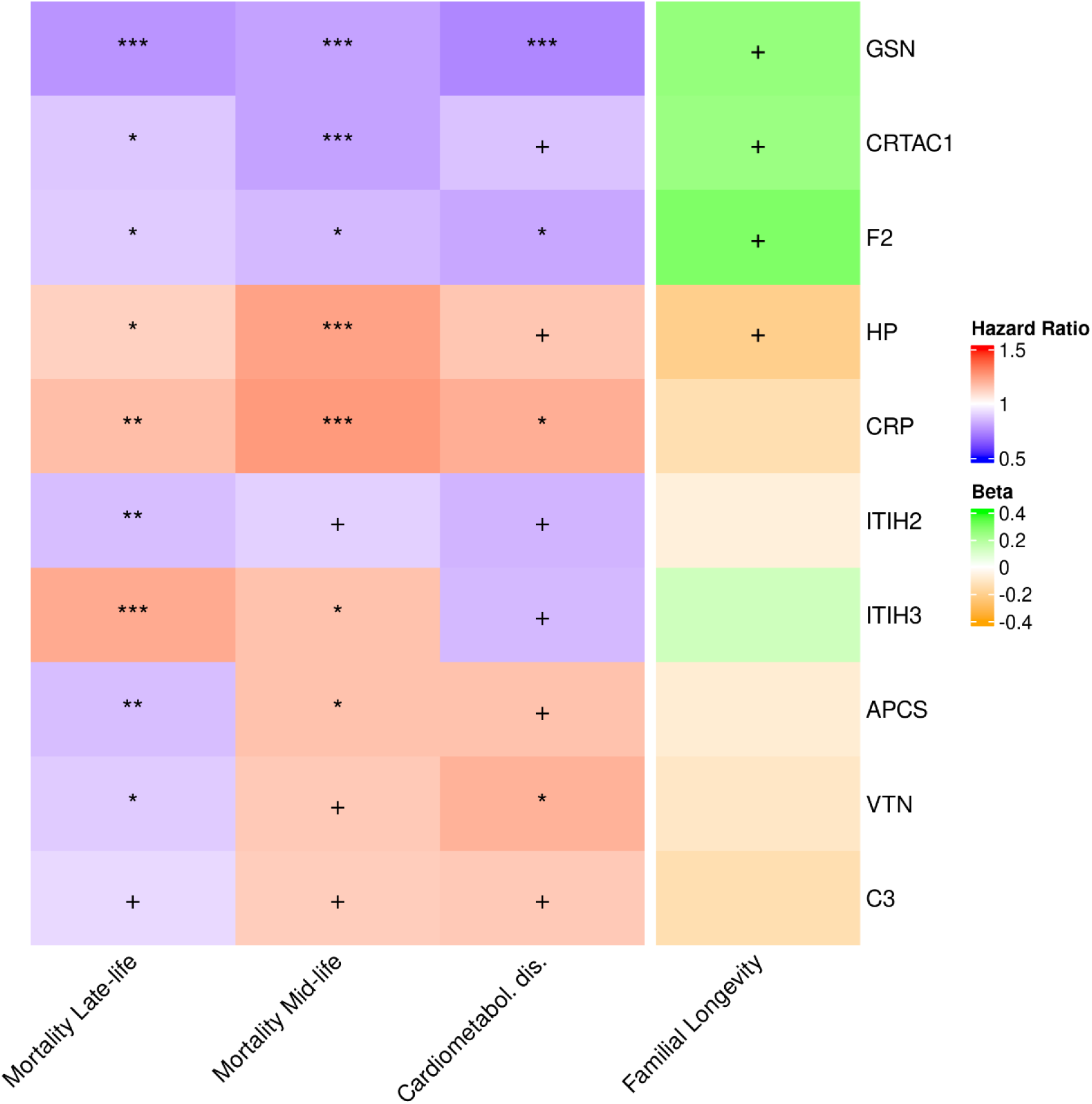
Candidate Protein Markers of Healthy Aging and Resilience. Heatmap showing a subgroup analysis of ten plasma proteins associated with survival, as defined as mortality, at mid- and late-life, and cardiometabolic healthspan, defined as cardiometabolic disease incidence, as well as their relationship with familial longevity. For the first three columns, cell color represents the hazard ratio, with increased risk shown in red and decreased risk in blue corresponding to higher protein levels in the blood. Only proteins significant in all three analyses (P ≤ 0.05) are included. The rightmost column shows the beta for associations with familial longevity, with green indicating an increase and orange a decrease in Longevity Relatives Count (LRC) score with higher protein levels. Significance in the cells is denoted as follows: P ≤ 0.05 = +, FDR ≤ 0.1 = *, FDR ≤ 0.01 = **, FDR ≤ 0.001 = ***.

In addition, consistent effect sizes were observed for CRP (higher protein levels linked to decreased survival) and ITIH2 (higher protein levels linked to increased survival) across all three outcomes, but these proteins did not associate with familial longevity. The remaining four proteins (ITIH3, APCS, VTN, and C3) showed mixed directional effects across survival in mid- and late-life and cardiometabolic healthspan.

Collectively these results suggest that GSN, CRTAC1, F2, and HP are promising candidate biomarkers of healthy aging and resilience, reflecting both longevity, increased survival and extended healthspan.

### Metabolome and proteome capture different biological mortality domains

Using ^1^H-NMR plasma metabolomics, we developed MetaboHealth, a biomarker of vulnerability (8). MetaboHealth significantly associated with mortality in both mid- and late-life in LLS (HR_F2_ 1.82 [95% CI: 1.47-2.27] and HR_F1_ 2.04 [95% CL: 1.80-2.30]). To determine if the mortality-associated proteins reflect distinct or overlapping biological mortality domains in comparison to MetaboHealth, we added MetaboHealth as a covariate to the proteomic survival models for both mid- and late-life.

We observed that 23 out of the 102 nominally significant proteins from the original late-life mortality analysis remained statistically significant after adding MetaboHealth to the model (**Supplementary Table 10**). Moreover, we observed that 24 out of the 49 proteins from the original mid-life mortality analysis remained statistically significant after adding MetaboHealth to the model (**Supplementary Table 11**). This indicates that they potentially capture independent survival features compared to MetaboHealth. When focusing on the 26 proteins that nominally significant overlapped between mid- and late-life mortality (**Figure 5**), we observed that six proteins (APCS, C7, FCN2, HPR, GSN, and PIGR; **Purple**) were significantly associated with survival, independent of MetaboHealth. Two of these proteins (GSN, and APCS) were also associated with cardiometabolic healthspan, further demarking their potential as health markers.

**Figure 5.**
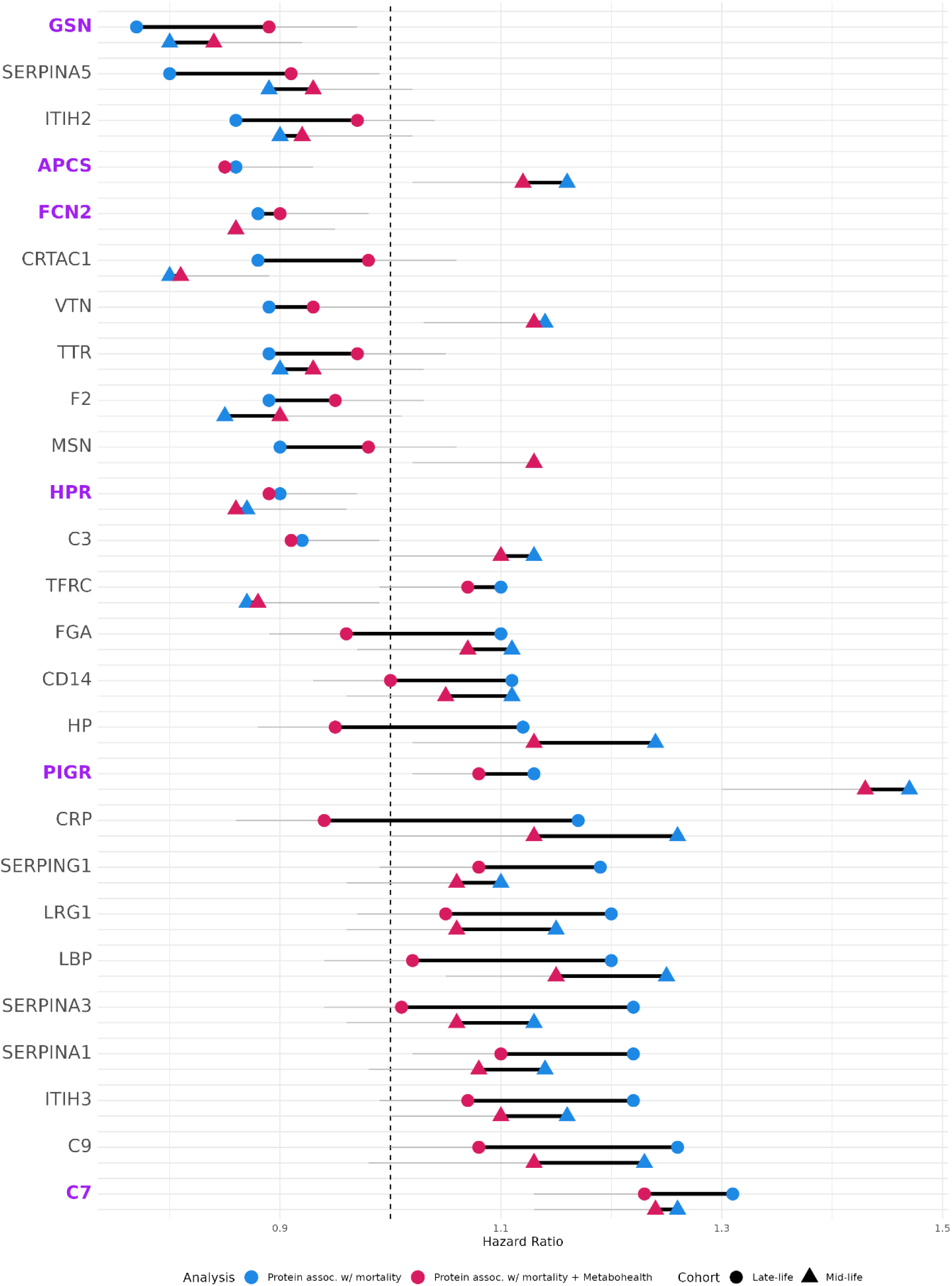
Sensitivity Analysis of Mortality Associated Proteins with marker of overall health MetaboHealth. Lollipop graph of shared proteins associated with mortality (P≤0.05; Blue; N=26) in both mid-life (Triangle) and late-life (Circle) and the sensitivity effect of previously developed ^1^H-NMR metabolomic-based marker of vulnerability MetaboHealth (Red). The shift between the two models indicates how much of the proteins mortality signal is also captured by the MetaboHealth biomarker. Outline indicated 95% confidence interval of the MetaboHealth analysis. Highlighted in purple and bold are the proteins that are both significant in its relation with mortality and independent of MetaboHealth in mid-life and late-life (N=6). Detailed results are presented in **Supplementary Table 7, 10, and 11**.

## Discussion

In this study, we used DIA-MS to measure the plasma proteome in the two generations of the family-based LLS and identified proteins potentially underlying healthy aging and resilience. We assessed the association between protein levels and long-term outcomes, including survival in mid- and late-life (up to 22 years follow-up) and cardiometabolic healthspan (up to 16 years follow-up). We identified four proteins, GSN, F2, HP, and CRTAC1, as candidate biomarkers of resilience and healthy aging. These proteins reflect resilience as they not only demonstrated consistent associations with survival across mid- and late-life, and cardiometabolic healthspan, but were also associated with the ancestral familial longevity score. We observed that higher levels of GSN, F2, and CRTAC1 associate with increased survival while higher levels of HP were linked to decreased survival and accelerated disease onset. Moreover, HP is less pronounced in long-lived families. In addition, we identified six proteins (APCS, C7, FCN2, HPR, GSN, and PIGR) that were significantly associated with survival, independent of MetaboHealth. Future studies could further investigate whether the addition of these proteins provides added value for health and survival prediction of MetaboHealth.

By analyzing data from two generations, we identified both novel and previously observed associations between proteins and survival that were age-dependent and consistent between mid- and late-life. Previous studies that investigated the association between proteins and survival mainly focused on disease cohorts (33), general population settings of middle-aged individuals (21, 22, 34-36), or older individuals (17, 18). Despite differences in protein measurement techniques and platforms many of our identified survival-associated proteins such as CRP (18, 21, 35), SERPINA3 (21, 34, 35), C7 (21, 34), and C9 (21, 34), were replicated and showed concordant associations across multiple studies (**Supplementary Table 12**). In comparison with the largest plasma proteome study on survival and age-related disease incidence utilizing the UK Biobank (22), only 3.5% (N=51/1468) of their analyzed proteins overlapped with the proteins we quantified. Nevertheless, 80% (N=12/15) of the shared survival-associated proteins (ITIH2, CRTAC1, CD14, PIGR, LBP, PON3, DPP4, CNDP1, PROC, NRP1, CST3, and AOC3) were replicated and directionally concordant (**Supplementary Table 13**).

Of the four potential biomarkers of resilience and healthy aging associated in our study three: gelsolin (GSN), coagulation factor II, prothrombin (F2), and haptoglobin (HP), are acute-phase proteins linked to inflammation of which the protein upregulation is largely driven by the hepatocytes (37). GSN, an actin-depolymerizing protein secreted largely by skeletal muscle (38), regulates immune responses and localizes inflammation (39), in line with our findings higher circulating levels were associated with increases survival (**Supplementary Table 12**; 18, 21). F2, a liver-derived proenzyme processed to thrombin (40), is essential for haemostasis and is a component measured in prothrombin time (PT) assay, a clinical measure of liver function and blood clotting. Higher F2 levels are linked to reduced paced of aging and have been implicated (along with CRP) as molecular readouts of social disadvantages in early- and late-life (41). HP is an IL-6 inducible protein (42), that binds free haemoglobin released from erythrocytes thereby preventing the loss of iron through the kidneys and bacterial uptake in case of infection, and is associated with higher levels with many inflammatory diseases, e.g. diabetes, cardiovascular disease, and obesity (42). Furthermore, Cartilage Acidic Protein 1, CRTAC1, a cartilage derived extracellular matrix protein (43), has emerged as a marker for osteoarthritis burden and progression (44–46), and is associated with increased survival, liver disease and diabetes onset (**Supplementary Table 14**; 22). Collectively, our findings, further underscore the central role of inflammation in healthy aging (47), and this aligns with the ‘inflammaging’ theory of aging,, whereby chronic systemic inflammation contributes to morbidity and early mortality (48,49).

MetaboHealth is further validated to be strongly associated with survival in both mid- and late-life, underscoring its value as a metabolomics-based biomarker for identifying individuals at increased risk of age-related health decline. In this study, we additionally identified six proteins (APCS, C7, FCN2, GSN, HPR, and PIGR) associated with survival independent of MetaboHealth in both mid- and late-life. With two of these proteins (GSN, and APCS) also being informative of healthspan in our analyses. While the metabolites underlying MetaboHealth are linked to lipid and fatty acid metabolism, glycolysis, fluid balance, and inflammation, the six proteins largely point towards the innate immune processes, i.e. inflammation, complement activation, and acute-phase responses, which partially overlap with the inflammatory pathways captured by Glycoprotein acetyls (GlycA) metabolite measurement in MetaboHealth. The GlycA test reflects systemic inflammation by capturing a composite signal from N-glycan side chains attached to several acute-phase proteins (such as CRP, HP, fibrinogen, and serum amyloid A), which undergo N-glycosylation during inflammatory responses (50). These findings suggest that integrating proteomic and metabolomic data could capture complementary aspects of inflammation and health and may improve biomarker-based prediction of survival and overall health (51).

There are some limitations to our study. First, our MS based panel focused on 326 proteins, largely representing the most abundant proteins in the blood. This means that previously identified important low-abundance markers associated with survival and morbidity, such as GDF-15 (34) or IL-6 (52), were not included in our analyses. However, the MS platform we used allows for high specificity and absolute quantification of the identified proteins, which would greatly facilitates further downstream biomarker discovery and assay development (53). With 33% (n=108) of the proteins in our panel being excluded from affinity-based platforms such as the Olink Explore 3k panel (2023), and SomaScan v4 platform (**Supplementary Table 14**). Although these proteins in our panel can be measured robustly, technical challenges persist as plasma MS proteomics still lacks full standardization, hindering consistent cross-study and cross-laboratory comparisons. Second, we assessed the protein-outcome relations based on a cross-sectional blood draw between two cohort in different life stages. While this study design allows for comparison across age groups, survival bias in the older generation, or other cohort selection effects, may influence the observed associations. Ideally these findings should be replicated in longitudinal cohorts, but cohorts with follow-up from middle to very old age are rare. Third, the LLS cohort consist of Dutch Caucasian individuals who are, on average healthier than the general population. As such, our findings may not directly generalize to more diverse or patient populations. Replication in non-Western and ethnically diverse groups is needed to assess the broader applicability of our candidate healthy aging markers. Finally, the analyses may be influenced by unmeasured social economic factors. However, the use of the family-based LLS design provides considerable control for such confounding. It is well-established that social economic factors are determinants of a wide range of adverse health outcomes (54), and as such proteomic and molecular readouts may also be used in future work to quantify social disadvantages in those at risk early on in life course (55).

In conclusion, we identified proteins significantly associated with long-term survival in mid-and late-life, cardiometabolic healthspan, and familial longevity. Our findings indicate that four proteins (GSN, F2, HP, and CRTAC1) capture multiple aspects of resilience, and we propose these as candidate biomarkers of healthy aging and resilience across the second half of the life course. These findings may give us better insights in the underlying mechanisms of healthy aging. Moreover, the development of proteomic biomarkers will pave the way for the construction of more sensitive indicators of health that can identify emerging health issues, before the onset of disease, thereby supporting earlier and more effective intervention, in both population health and clinical care.

## Supporting information

Supplementary Information Longevity Proteomics

## Data Availability

The datasets analyzed in the current study are available following a data access procedure (https://leidenlangleven.nl/data-access/). For each request it will be tested whether the research is in compliance with the informed consent that has been signed by the LLS participants.
This study does not report original code. All data were analysed and processed using published software packages, the details of which are provided and cited in the Methods section.

## Acknowledgements

We would like to thank Mar Rodríguez-Girondo for her valuable statistical input, and Fatih Bogaards for his support with the proteomic data. We are also grateful to Eka H D Suchiman for her assistance with the sample preparation. The contribution of all participants in the Leiden Longevity Study is gratefully acknowledged; this research would not have been possible without their involvement.

## Contributions

PCP, MB, JD, PES and, NvdB conceived the study and interpreted the data. PES and NvdB acquired funding and jointly supervised the project. NL, JWL, SM, RUM, and PA generated the data. PCP performed the formal analysis, validation of the data, and created the visualizations. PCP, NvdB and JD wrote the initial draft of the manuscript, with additional contributions from PES, MB, and PA. All authors provided critical feedback and approved the final manuscript.

## Data sharing statement

The datasets analyzed in the current study are available following a data access procedure (https://leidenlangleven.nl/data-access/). For each request it will be tested whether the research is in compliance with the informed consent that has been signed by the LLS participants.

This study does not report original code. All data were analysed and processed using published software packages, the details of which are provided and cited in the Methods section.

## Abbreviations

AcAce: Acetoacetate
Alb: Albumin
AP: Angina pectoris
CMD: Cardiometabolic disease
CVA: Cerebrovascular accident
DIA: Data independent acquisition
FDR: False Discovery Rate
Glc: Glucose
GP: General Practitioner
Gp / GlycA: Glycoprotein acetyls
His: Histidine
ICD-10: International Statistical Classification of Diseases and Related Health Problems
Ile: Isoleucine
Lac: Lactate
Leu: Leucine
LLS: Leiden Longevity Study
LMM: Linear Mixed Models
LRC-score: Longevity Relatives Count score
MI: Myocardial infarction
MS: Mass spectrometry
NMR: Nuclear magnetic resonance
Phe: Phenylalanine
PRD: Personal Records Database
PT: Prothrombin time
PUFA/FA: Ratio of polyunsaturated fatty acids to total fatty acids
RIN: Rank-Inversed Normalized
S-HDL-L: Total lipids in small HDL
TIA: Transient Ischemic Attack
Val: Valine
VLDL-D: mean diameter for VLDL particles
XXL-VLDL-L: Total lipids in chylomicrons and extremely large VLDL

## References

1. Barnett K, Mercer SW, Norbury M, Watt G, Wyke S, Guthrie B. Epidemiology of multimorbidity and implications for health care, research, and medical education: a cross-sectional study. Lancet. 2012;380(9836):37–43.

2. Ferrari AJ, Santomauro DF, Aali A, Abate YH, Abbafati C, Abbastabar H, et al. Global incidence, prevalence, years lived with disability (YLDs), disability-adjusted life-years (DALYs), and healthy life expectancy (HALE) for 371 diseases and injuries in 204 countries and territories and 811 subnational locations, 1990-2021: a systematic analysis for the Global Burden of Disease Study 2021. Lancet. 2024;403(10440):2133–61.

3. Kennedy BK, Berger SL, Brunet A, Campisi J, Cuervo AM, Epel ES, et al. Geroscience: Linking Aging to Chronic Disease. Cell. 2014;159(4):708–12.

4. Partridge L, Deelen J, Slagboom PE. Facing up to the global challenges of ageing. Nature. 2018;561(7721):45–56.

5. Kuiper LM, Polinder-Bos HA, Bizzarri D, Vojinovic D, Vallerga CL, Beekman M, et al. Epigenetic and Metabolomic Biomarkers for Biological Age: A Comparative Analysis of Mortality and Frailty Risk. J Gerontol A Biol Sci Med Sci. 2023;78(10):1753–62.

6. Peters MJ, Joehanes R, Pilling LC, Schurmann C, Conneely KN, Powell J, et al. The transcriptional landscape of age in human peripheral blood. Nat Commun. 2015;6:8570.

7. Oh HS, Rutledge J, Nachun D, Palovics R, Abiose O, Moran-Losada P, et al. Organ aging signatures in the plasma proteome track health and disease. Nature. 2023;624(7990):164–72.

8. Deelen J, Kettunen J, Fischer K, van der Spek A, Trompet S, Kastenmüller G, et al. A metabolic profile of all-cause mortality risk identified in an observational study of 44,168 individuals. Nature Communications. 2019;10.

9. Zonneveld MH, Al Kuhaili N, Mooijaart SP, Slagboom PE, Jukema JW, Noordam R, et al. Increased H-NMR metabolomics-based health score associates with declined cognitive performance and functional independence in older adults at risk of cardiovascular disease. Geroscience. 2024.

10. Venema JA, Kuranova A, Bizzarri D, Mooijaart SP, Kerckhoffs APM, Slieker K, et al. Associations between metabolomic scores and clinical outcomes in hospitalized COVID-19 patients. Geroscience. 2025.

11. Anderson NL, Anderson NG. The human plasma proteome - History, character, and diagnostic prospects. Mol Cell Proteomics. 2002;1(11):845–67.

12. Lehallier B, Gate D, Schaum N, Nanasi T, Lee SE, Yousef H, et al. Undulating changes in human plasma proteome profiles across the lifespan. Nat Med. 2019;25(12):1843–50.

13. Tanaka T, Biancotto A, Moaddel R, Moore AZ, Gonzalez-Freire M, Aon MA, et al. Plasma proteomic signature of age in healthy humans. Aging Cell. 2018;17(5).

14. Menni C, Kiddle SJ, Mangino M, Vinuela A, Psatha M, Steves C, et al. Circulating Proteomic Signatures of Chronological Age. J Gerontol A Biol Sci Med Sci. 2015;70(7):809–16.

15. Sun BB, Chiou J, Traylor M, Benner C, Hsu YH, Richardson TG, et al. Plasma proteomic associations with genetics and health in the UK Biobank. Nature. 2023;622(7982):329–38.

16. Coenen L, Lehallier B, de Vries HE, Middeldorp J. Markers of aging: Unsupervised integrated analyses of the human plasma proteome. Front Aging. 2023;4:1112109.

17. Sathyan S, Ayers E, Gao TN, Weiss EF, Milman S, Verghese J, et al. Plasma proteomic profile of age, health span, and all-cause mortality in older adults. Aging Cell. 2020;19(11).

18. Orwoll ES, Wiedrick J, Jacobs J, Baker ES, Piehowski P, Petyuk V, et al. High-throughput serum proteomics for the identification of protein biomarkers of mortality in older men. Aging Cell. 2018;17(2).

19. Hantikainen E, Weichenberger CX, Dordevic N, Hernandes VV, Foco L, Goegele M, et al. Metabolite and protein associations with general health in the population-based CHRIS study. Sci Rep-Uk. 2024;14(1).

20. Liu XJ, Axelsson GT, Newman AB, Psaty BM, Boudreau RM, Wu CK, et al. Plasma proteomic signature of human longevity. Aging Cell. 2024;23(6).

21. Tanaka T, Basisty N, Fantoni G, Candia J, Moore AZ, Biancotto A, et al. Plasma proteomic biomarker signature of age predicts health and life span. Elife. 2020;9.

22. Gadd DA, Hillary RF, Kuncheva Z, Mangelis T, Cheng YP, Dissanayake M, et al. Blood protein assessment of leading incident diseases and mortality in the UK Biobank. Nature Aging. 2024;4(8).

23. van den Berg N, Rodriguez-Girondo M, van Dijk IK, Mourits RJ, Mandemakers K, Janssens A, et al. Longevity defined as top 10% survivors and beyond is transmitted as a quantitative genetic trait. Nat Commun. 2019;10(1):35.

24. van den Berg N, Rodriguez-Girondo M, van Dijk IK, Slagboom PE, Beekman M. Increasing number of long-lived ancestors marks a decade of healthspan extension and healthier metabolomics profiles. Nature Communications. 2023;14(1).

25. Schoenmaker M, de Craen AJ, de Meijer PH, Beekman M, Blauw GJ, Slagboom PE, et al. Evidence of genetic enrichment for exceptional survival using a family approach: the Leiden Longevity Study. Eur J Hum Genet. 2006;14(1):79–84.

26. van den Berg N, Rodriguez-Girondo M, Mandemakers K, Janssens AAPO, Beekman M, Slagboom PE. Longevity Relatives Count score identifies heritable longevity carriers and suggests case improvement in genetic studies. Aging Cell. 2020;19(6).

27. Van der Meulen A. Life tables and Survival analysis. 2012.

28. Hughes CS, Moggridge S, Müller T, Sorensen PH, Morin GB, Krijgsveld J. Single-pot, solid-phase-enhanced sample preparation for proteomics experiments. Nat Protoc. 2019;14(1):68–+.

29. Rappsilber J, Mann M, Ishihama Y. Protocol for micro-purification, enrichment, pre-fractionation and storage of peptides for proteomics using StageTips. Nat Protoc. 2007;2(8):1896–906.

30. Chambers MC, Maclean B, Burke R, Amodei D, Ruderman DL, Neumann S, et al. A cross-platform toolkit for mass spectrometry and proteomics. Nat Biotechnol. 2012;30(10):918–20.

31. Demichev V, Messner CB, Vernardis SI, Lilley KS, Ralser M. DIA-NN: neural networks and interference correction enable deep proteome coverage in high throughput. Nat Methods. 2020;17(1):41–+.

32. Balan TA, Putter H. frailtyEM: An R Package for Estimating Semiparametric Shared Frailty Models. J Stat Softw. 2019;90(7).

33. Luo H, Petrera A, Hauck SM, Rathmann W, Herder C, Gieger C, et al. Association of plasma proteomics with mortality in individuals with and without type 2 diabetes: Results from two population-based KORA cohort studies. Bmc Med. 2024;22(1).

34. Eiriksdottir T, Ardal S, Jonsson BA, Lund SH, Ivarsdottir EV, Norland K, et al. Predicting the probability of death using proteomics. Commun Biol. 2021;4(1):758.

35. Ho JE, Lyass A, Courchesne P, Chen G, Liu CY, Yin XY, et al. Protein Biomarkers of Cardiovascular Disease and Mortality in the Community. J Am Heart Assoc. 2018;7(14).

36. Molvin J, Jujic A, Melander O, Pareek M, Råstam L, Lindblad U, et al. Proteomic exploration of common pathophysiological pathways in diabetes and cardiovascular disease. Esc Heart Fail. 2020;7(6):4151–8.

37. Gabay C, Kushner I. Mechanisms of disease: Acute-phase proteins and other systemic responses to inflammation. New Engl J Med. 1999;340(6):448–54.

38. Kwiatkowski DJ, Mehl R, Izumo S, Nadalginard B, Yin HL. Muscle Is the Major Source of Plasma Gelsolin. J Biol Chem. 1988;263(17):8239–43.

39. Li GH, Arora PD, Chen Y, McCulloch CA, Liu P. Multifunctional roles of gelsolin in health and diseases. Med Res Rev. 2012;32(5):999–1025.

40. Lane DA, Philippou H, Huntington JA. Directing thrombin. Blood. 2005;106(8):2605–12.

41. Kivimaki M, Pentti J, Frank P, Liu F, Blake A, Nyberg ST, et al. Social disadvantage accelerates aging. Nat Med. 2025.

42. Quaye IK. Haptoglobin, inflammation and disease. T Roy Soc Trop Med H. 2008;102(8):735–42.

43. Steck E, Bräun J, Pelttari K, Kadel S, Kalbacher H, Richter W. Chondrocyte secreted CRTAC1:: A glycosylated extracellular matrix molecule of human articular cartilage. Matrix Biology. 2007;26(1):30–41.

44. Styrkarsdottir U, Lund SH, Saevarsdottir S, Magnusson M, Gunnarsdottir K, Norddahl GL, et al. The CRTAC1 Protein in Plasma Is Associated With Osteoarthritis and Predicts Progression to Joint Replacement: A Large-Scale Proteomics Scan in Iceland. Arthritis Rheumatol. 2021;73(11):2025–34.

45. Szilagyi IA, Vallerga CL, Boer CG, Schiphof D, Ikram MA, Bierma-Zeinstra SMA, et al. Plasma proteomics identifies CRTAC1 as a biomarker for osteoarthritis severity and progression. Rheumatology. 2023;62(3):1286–95.

46. Styrkarsdottir U, Lund SH, Thorleifsson G, Saevarsdottir S, Gudbjartsson DF, Thorsteinsdottir U, et al. Cartilage Acidic Protein 1 in Plasma Associates With Prevalent Osteoarthritis and Predicts Future Risk as Well as Progression to Joint Replacements: Results From the UK Biobank Resource. Arthritis Rheumatol. 2023;75(4):544–52.

47. Haynes L. Aging of the Immune System: Research Challenges to Enhance the Health Span of Older Adults. Front Aging-Lausanne. 2020;1.

48. Proctor MJ, McMillan DC, Horgan PG, Fletcher CD, Talwar D, Morrison DS. Systemic Inflammation Predicts All-Cause Mortality: A Glasgow Inflammation Outcome Study. Plos One. 2015;10(3).

49. Ferrucci L, Fabbri E. Inflammageing: chronic inflammation in ageing, cardiovascular disease, and frailty. Nat Rev Cardiol. 2018;15(9):505–22.

50. Ballout RA, Remaley AT. GlycA: A New Biomarker for Systemic Inflammation and Cardiovascular Disease (CVD) Risk Assessment. J Lab Precis Med. 2020;5.

51. Sun YV, Hu YJ. Integrative Analysis of Multi-omics Data for Discovery and Functional Studies of Complex Human Diseases. Adv Genet. 2016;93:147–90.

52. Tylutka A, Walas L, Zembron-Lacny A. Level of IL-6, TNF, and IL-1beta and age-related diseases: a systematic review and meta-analysis. Front Immunol. 2024;15:1330386.

53. Hartl J, Kurth F, Kappert K, Horst D, Mulleder M, Hartmann G, et al. Quantitative protein biomarker panels: a path to improved clinical practice through proteomics. EMBO Mol Med. 2023;15(4):e16061.

54. Kivimäki M, Batty GD, Pentti J, Shipley MJ, Sipilä PN, Nyberg ST, et al. Association between socioeconomic status and the development of mental and physical health conditions in adulthood: a multi-cohort study. Lancet Public Health. 2020;5(3):E140–E9.

55. Potente C. Proteomics sheds light on unequal aging. Nat Med. 2025.

